# Substantial underestimation of SARS-CoV-2 infection in the United States due to incomplete testing and imperfect test accuracy

**DOI:** 10.1101/2020.05.12.20091744

**Authors:** Sean L. Wu, Andrew Mertens, Yoshika S. Crider, Anna Nguyen, Nolan N. Pokpongkiat, Stephanie Djajadi, Anmol Seth, Michelle S. Hsiang, John M. Colford, Art Reingold, Benjamin F. Arnold, Alan Hubbard, Jade Benjamin-Chung

## Abstract

Accurate estimates of the burden of SARS-CoV-2 infection are critical to informing pandemic response. Current confirmed COVID-19 case counts in the U.S. do not capture the total burden of the pandemic because testing has been primarily restricted to individuals with moderate to severe symptoms due to limited test availability. We used a semi-Bayesian method to perform a probabilistic bias analysis on cumulative confirmed COVID-19 counts, incorporating relevant prior knowledge from existing studies about SARS-CoV-2 testing probabilities and diagnostic accuracy parameters while accounting for uncertainty. We estimate 6,275,072 cumulative infections compared to 721,245 confirmed cases (1.9% vs. 0.2% of the population) as of April 18, 2020. Accounting for uncertainty, the number of infections was 3 to 20 times higher than the number of confirmed cases. 86% (simulation interval: 64-99%) of this difference was due to incomplete testing, while 14% (0.3-36%) was due to imperfect test accuracy. Estimates of SARS-CoV-2 infections that transparently account for testing practices and diagnostic accuracy reveal that the pandemic is larger than confirmed case counts suggest.

---

The severe acute respiratory syndrome-coronavirus 2 (SARS-CoV-2) pandemic is reported to have caused 721,245 confirmed cases of coronavirus disease 2019 (COVID-19) in the U.S. by April 18, 2020. The first known case in the U.S. was confirmed on January 21, 2020. In February, SARS-CoV-2 testing remained limited due to flawed test kits. As of mid-April, the U.S. Centers for Disease Control and Prevention (CDC) recommended that physicians prioritize testing hospitalized patients, who tend to have moderate to severe symptoms. Most state testing policies were consistent with this recommendation (Appendix Table 1). Yet, evidence from studies that conducted broader testing suggest that 30-70% of individuals who test positive have mild or no symptoms^2-6^ and that asymptomatic and pre-symptomatic individuals can transmit SARS-CoV-2.^7-9^ Thus, a substantial number of mild or asymptomatic infections in the U.S. may be undetected. Furthermore, initial evidence suggests that tests based on nasopharyngeal and throat swabs may produce false negative results.^10-13^ Thus, counts of confirmed cases are biased due to incomplete testing and imperfect test sensitivity. Accurate estimates of the burden of SARS-CoV-2 infection are critical to understanding the course of the pandemic and informing public health response.

To date, the majority of studies that have estimated the burden of SARS-CoV-2 infection have used mathematical models (e.g., compartmental or agent-based models).^14-18^ Mathematical modeling studies attempt to mimic natural disease transmission systems by modeling age and social structure, travel and commuting patterns, and immune dynamics. Such models are particularly useful for projecting transmission patterns under hypothetical interventions but can be highly complex; when sufficient data do not exist to parameterize such models, results can be quite biased.^19,20^ In addition, model outputs are sensitive to assumed population structure and contact patterns, which are difficult to validate, particularly in a novel pathogen setting.

We estimated the total number of SARS-CoV-2 infections in each U.S. state from February 28 to April 18, 2020 using probabilistic bias analysis, a semi-Bayesian approach, to correct empirical confirmed case counts for bias due to incomplete testing and imperfect test accuracy. This method is commonly used to quantify the impact of and correct for measurement bias in epidemiologic studies.^21^ Our objective was to estimate infection burden from empirical data rather than forecast future dynamics. While our approach and mathematical models both require difficult to verify assumptions for which little evidence is currently available, the simplicity of our approach facilitates transparent assessment of model assumptions.

Using the available evidence, we defined prior distributions of testing probabilities among individuals with moderate to severe symptoms requiring medical attention or hospitalization (e.g., shortness of breath, high fever) vs. those with minimal symptoms (e.g., cough without difficulty breathing or shortness of break, low grade fever) or no symptoms (Table 1, Appendix Figure 1). The Methods section includes a detailed description for our assumed prior distributions. To quantify uncertainty in testing probabilities, we randomly sampled from each prior distribution and estimated the total number of infections using a simple model relating the number of individuals tested to testing probabilities. We then repeated this process 1,000 times to obtain a distribution of estimated SARS-CoV-2 infections in each state. We report the medians of the distribution of infections in each state and the simulation interval (2.5^th^ and 97.5^th^ percentiles). The estimated number of infections included confirmed COVID-19 cases and undiagnosed infections among individuals mild or no symptoms.

**Table 1.** Prior distributions for probabilistic bias analysis

| **Distribution** | **Minimum** | **Median** | **Maximum** |
| --- | --- | --- | --- |
| $P(S_1 \vert \text{tested})$ | 0.557 | 0.947 | 1.000 |
| $P(S_1 \vert \text{untested})$ | 0.002 | 0.052 | 0.149 |
| $\alpha$ | 0.800 | 0.909 | 0.988 |
| $\beta$ | 0.003 | 0.046 | 0.318 |
| $P(\underline{S}_0 \vert \text{test } +)$ | 0.251 | 0.488 | 0.700 |
| Sensitivity | 0.650 | 0.872 | 1.000 |
| Specificity | 0.9998 | 0.999 | 1.000 |

Detailed descriptions of each prior distribution are included in the Methods section.

We defined a prior distribution for the probability of being symptomatic among those tested, ranging from 0.6 to 1.0 under the assumption that the vast majority of individuals tested by mid-April in the U.S. were hospitalized and/or moderately to severely ill. We assumed that the probability of being symptomatic among untested individuals ranged from 0.002 to 0.149. In each state, we estimated the probability that untested individuals would have tested positive had they been tested. For those with moderate to severe symptoms, we assumed that the probability of testing positive was 80% to 99% the empirical proportion of tests that were positive in each state. For those who had mild or no symptoms, we assumed that the probability of testing positive was 0.2% to 32% the empirical proportion of tests that were positive in each state.

We also defined a prior for the probability of having minimal or no symptoms among those who tested positive; because five studies published estimates of this probability,^2-6^ we considered this prior to be more reliable than others. We thus used Bayesian melding to constrain the other prior distributions such that the probability of having minimal or no symptoms among those who tested positive ranged from 0.25 to 0.70, consistent with published studies.^22^ Priors were the same across states except for the probability of testing positive conditional on symptom status, which was a function of the empirical proportion of tests that were positive in each state (Appendix Table 2).

We quantified the SARS-CoV-2 testing rate per 1,000 using daily counts of tests in each state and 2019 projected state populations from the 2010 U.S. Census.^23^ By April 18, 2020, the SARS-CoV-2 testing rate was 11 per 1,000 in the U.S. However, there were large discrepancies in testing between states, with state-level testing rates of 6 per 1,000 in Kansas to 31 per 1,000 in Rhode Island (Figure 1). Generally, testing rates were higher in the Northwest and Northeast and lower in the Midwest and South.

**Figure 1.**
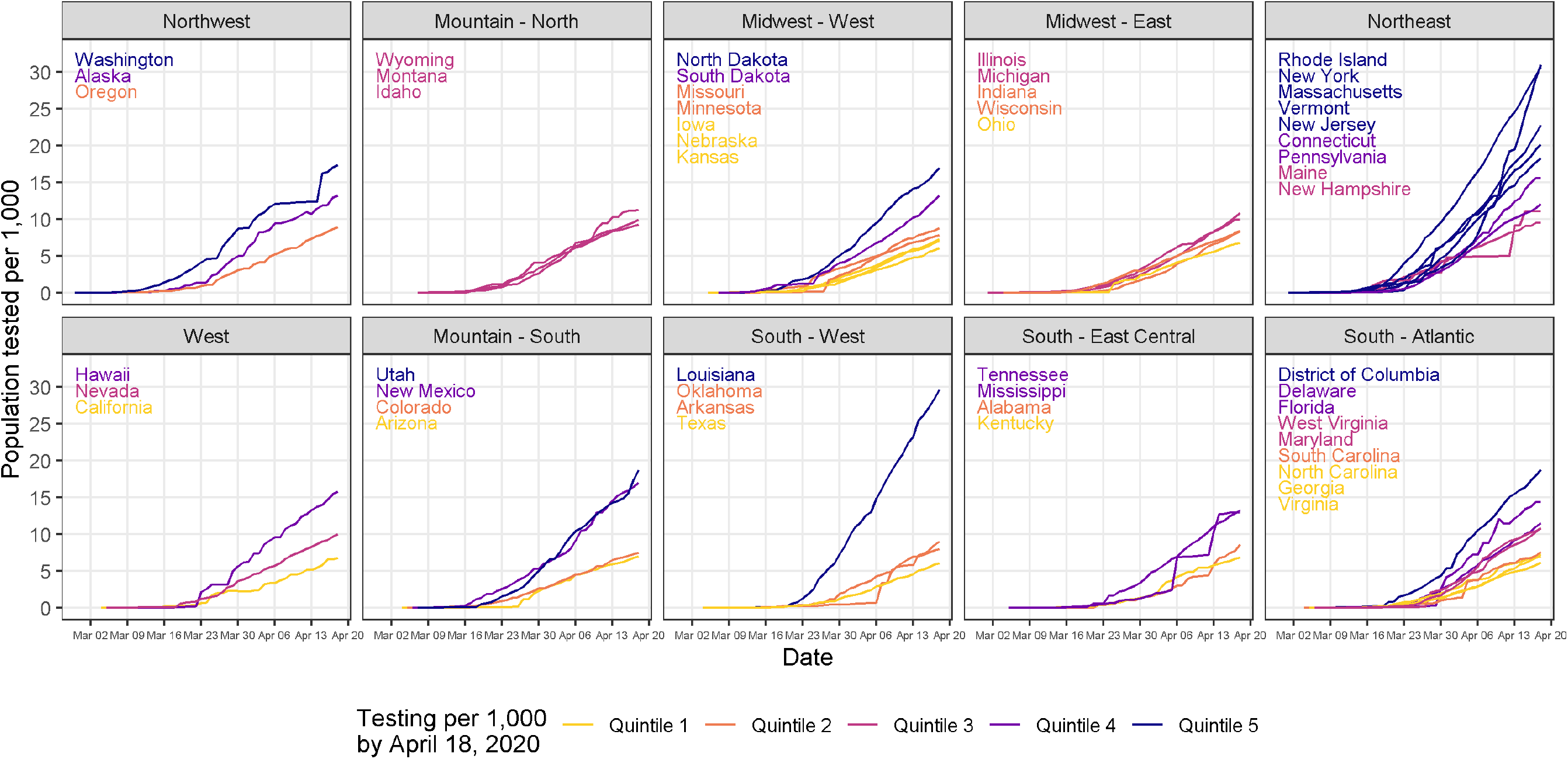
Daily SARS-CoV-2 tests per 1,000 population in the United States by state and region.

When correcting for incomplete testing and imperfect test accuracy, we estimated that the total number of SARS-CoV-2 infections in the U.S. by April 18, 2020 was 6,275,072 (19 per 1,000) – an estimate 9 times larger than the 721,245 confirmed cases (2 per 1,000) reported during this period. These results imply that 89% of infections in the US were undocumented. This finding is consistent with a mathematical modeling study that reported that 86% of infections were undocumented using data from Wuhan, China.^17^ The simulation interval for the number of estimated infections in the U.S. was 2,193,224 to 14,187,611 (Appendix Figure 2). This corresponds to an estimated number of SARS-CoV-2 infections 3 to 20 times higher than the number of confirmed cases. Nationally, we estimate that 86% (simulation interval: 64-99%) of the difference between confirmed cases and estimated infections was due to incomplete testing, and 14% (simulation interval: 0.3-36%) of the difference was due to imperfect test accuracy.

Disparities between confirmed SARS-CoV-2 infections and estimated total infections varied widely by state and geographic region. In each state, confirmed COVID-19 case counts ranged from 0.4 to 12.2 per 1,000, while estimated total infections ranged from 3.0 to 63.0 per 1,000 (Figure 2a, Appendix Figure 2). Compared to confirmed COVID-19 case counts, expected infections were 5 to 19 times larger (Figure 2b). COVID-19 incidence was highest in the northeast, Midwest, and Louisiana using confirmed case counts (Figure 3A) or estimated infections (Figure 2A). However, underestimation of SARS-CoV-2 infections was more common in California, the Midwest, and some Southern states (Figure 3B). In 32 states, the number of infections was at least 10 times larger than the number of confirmed cases. Differences in state-specific results are driven by observed differences in transmission, testing rates, and test positivity rates in each state rather than our modeling assumptions. In states with the largest underestimates of SARS-CoV-2 infections, public health responses based on confirmed cases may be inadequate to reduce transmission.

**Figure 2.**
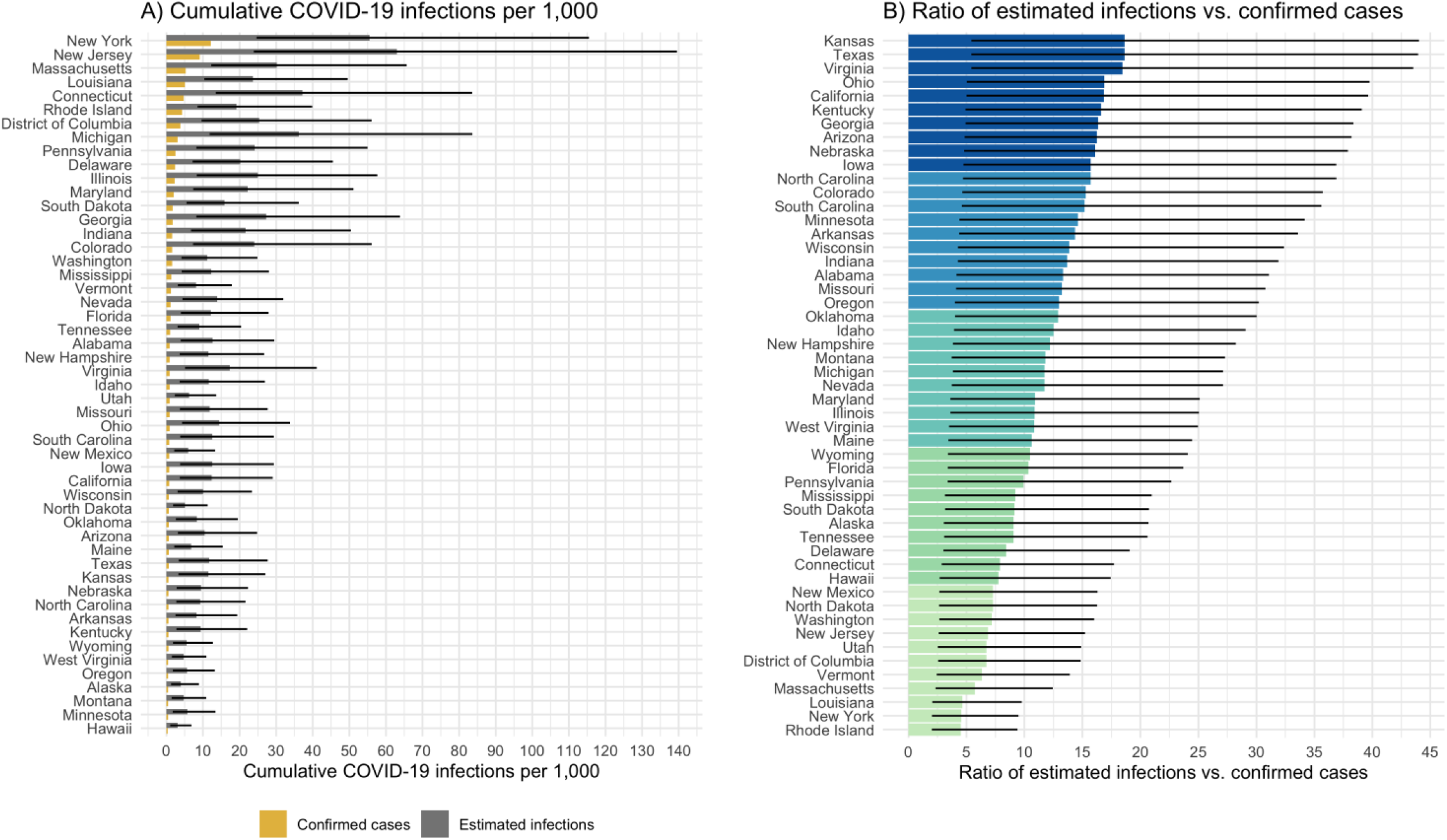
Cumulative confirmed COVID-19 cases vs. estimated SARS-CoV-2 infections correcting for incomplete testing and imperfect test accuracy in the United States.

**Figure 3.**
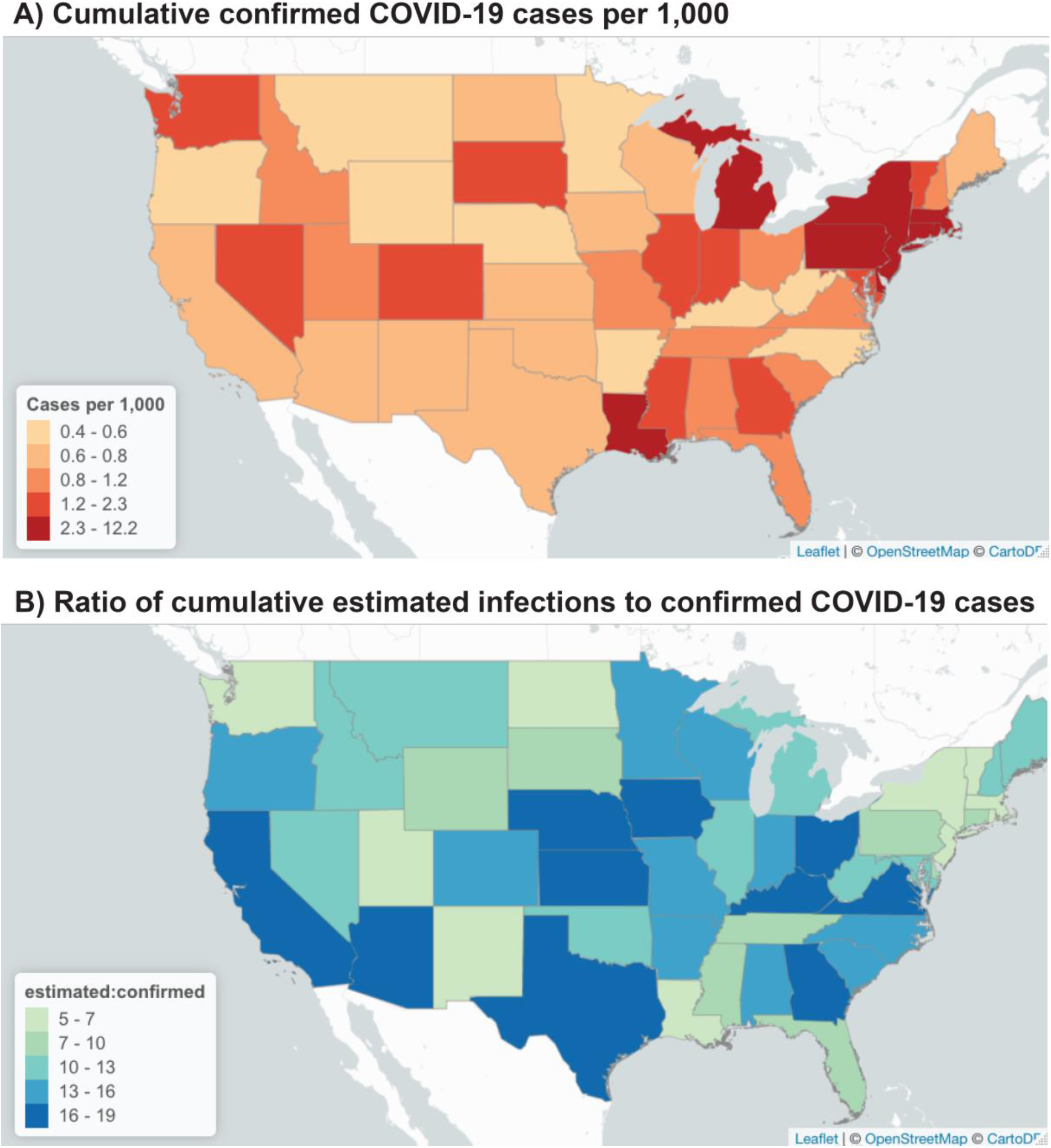
Map of cumulative confirmed COVID-19 case counts and the ratio of expected infections correcting for bias due to incomplete testing and imperfect test accuracy to confirmed cases in the United States.

Our findings reveal underestimation of the complete burden of the SARS-CoV-2 pandemic in the U.S. We have illustrated the importance of adjusting estimates of COVID-19 infections for testing rates and diagnostic accuracy during a period of low testing rates. A strength of our approach is that it quantifies the contribution of incomplete testing vs. imperfect test accuracy to under-estimation of the burden of COVID-19, demonstrating that the majority was due to incomplete testing. Our methods are not specific to the diagnostic test used and can be applied to serological studies to account for the imperfect diagnostic accuracy of serological tests and non-random sampling strategies used in some serological studies.

Little is known about whether infection with SARS-CoV-2 confers lasting immunity, and if so, for how long. Even in a best-case scenario in which SARS-CoV-2 infection produces immunity for one to two years, as is common for other betacoronaviruses,^24^ our results suggest that a very small proportion of the population has developed immunity and that the U.S. is not close to achieving herd immunity.

Our findings are broadly consistent with other studies using different methods to estimate the total SARS-CoV-2 infections. A mathematical modeling study projected 560 cumulative infections per 1,000 in the U.S. in 2020 if social distancing had been implemented population-wide.^15^ In comparison, we estimated 19 infections per 1,000 over a 25-day period in an early stage of the pandemic prior to peak transmission. Other studies from countries where time-indexed district-level hospitalization and mortality data are readily available estimated the proportion of the population infected with SARS-CoV-2 in Italy and France, where testing rates are higher.^25,26^ These studies estimate that the number of infections is at least twice the number of confirmed cases, consistent with our findings.

A limitation of this study is that our prior distributions are based on limited initial evidence about SARS-CoV-2 testing probabilities. Though we used the best available evidence at this early stage of the pandemic, it is possible that our priors do not reflect true testing probabilities. For example, our testing priors were informed by state-level testing guidelines, which typically prioritize groups for testing and allow physician discretion in ordering tests. In late March, at least eight states expanded their testing eligibility criteria to include outpatients with mild symptoms, and at least four states expanded eligibility to include asymptomatic individuals who had close contact with a confirmed case (Appendix Table 1). These changes may imply different priors in these states, however, severe test supply shortages have been widely documented in news reports and in state-level guidance to health professionals, suggesting that testing of low priority groups during this study was minimal.

Another limitation is that our model used state-specific estimates of the probability of testing positive among individuals who were tested. In states with very low testing rates, empirical test positive probabilities may not accurately reflect incidence in the general population due to prioritized testing of the severely ill and special populations. Nevertheless, our wide simulation intervals reflect these uncertainties in our prior distributions. Even using the lower bound of the simulation interval for the U.S. as a conservative estimate suggests that confirmed case counts underestimate total infections by a factor of 3. Results from future studies that rigorously estimate the incidence of symptomatic and asymptomatic SARS-CoV-2 infection can be used to update our priors and improve the precision and accuracy of our estimates.

State-level data on SARS-CoV-2 positive tests may mask meaningful geographic variation at a smaller scale. Unfortunately, county-level estimates of the number of individuals tested were not readily available at the time of this study. In addition, our model did not account for state-specific variation in data quality due to incomplete reporting of testing in some states (Appendix Table 3).

We did not estimate the infection fatality ratio because COVID-19 deaths outside hospitals are likely to be underreported, and death registrations may not be up to date. COVID-19-specific death reporting rates likely differ between tested and untested individuals, but the extent of this difference is difficult to quantify. In addition, accurate estimation of the infection fatality ratio would account for age, but age-stratified counts of COVID-19 deaths in each state are not readily available.

Underestimates of the number of SARS-CoV-2 infections jeopardize the success of pandemic response policies: they suggest to the public that the threat of the pandemic is smaller than it is, making it difficult to justify stringent social distancing policies. Our results highlight the urgent need to systematically expand SARS-CoV-2 testing across the U.S. to provide an accurate evidence base for pandemic response policies.

## Methods

### Data

We obtained 2019 projected state populations (N) from the 2010 U.S. Census and observed daily counts of tests (*N*_tested_) and confirmed SARS-CoV-2 positive tests 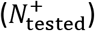 in each state from February 28 to April 18, 2020. Data was collected by the COVID Tracking Project,^23^ which assembles data on a regular basis primarily from state, district, and territory public health departments. National case counts from this source were comparable with those from the CDC on April 18 (COVID Tracking Project: 721,245; CDC: 720,630). State-level reporting of tests completed and the number of positive tests by date varied; COVID Tracking Project assigned each state a data quality grade based on whether 1) reporting positives reliably, 2) reporting negatives sometimes, 3) reporting negatives reliably, and 4) reporting all commercial tests. Thirty-five states and the District of Columbia met all of these criteria, 14 states met three criteria, and one state met only two criteria (Appendix Table 3). We assumed that all test results included in this data source were done using polymerase chain reaction because during the study period, alternative tests (e.g., antibody tests) were not approved for diagnosis of SARS-CoV-2 infection by the U.S. Food and Drug Administration (such tests were only used for research purposes and are not likely to have been included in case counts). In addition, we assumed that the vast majority of samples collected were nasopharyngeal swabs, which were recommended by the CDC as the preferred choice of swab.^27^

### Correction for incomplete testing

To correct for incomplete testing, we defined the following formulas to estimate the number of SARS-CoV-2 infections among individuals who were not tested had they been tested 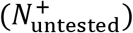. Below, *S_1_* is an indicator for having moderate to severe COVID-19 symptoms, and *S_0_* is an indicator for having minimal to no COVID-19 symptoms.

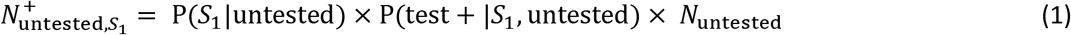

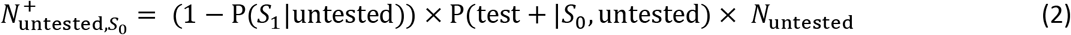

where P(*S_1_* |tested) is the probability of being symptomatic if tested, and P(*S_1_* |untested) is the probability of being symptomatic if not tested, P(test + | *S_1_*, untested) is the probability of testing positive among the untested who had moderate to severe symptoms had they been tested, and P(test +1 *S_0_*, untested) is the analogous probability among the untested who had mild or no symptoms.

Equations 1 and 2 can be interpreted probabilistically by noting that *N*_untested_ is proportional to P(untested), the probability that a random individual in the population has not been tested. Writing 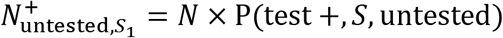 and *N*_untested_ = *N*× P(untested), where *N* is the total population size, dividing by *N* we are left with P(test +,*S_1_*, untested) = P(test + |*S_1_*,untested) × P(*S_1_*|untested) × P(untested), and likewise for Equation 2. Thus, we can reinterpret these equations as giving the joint probability that an untested individual with moderate or severe symptoms (mild or no symptoms) tested positive factorized as the marginal probability of being untested multiplied by the two conditional probabilities. Scaling by population size (interpreting probabilities as proportions) we arrive at Equations 1 and 2.

For untested individuals who had moderate to severe symptoms, we assumed that the probability they tested positive was 80% to 99% the empirical probability of testing positive among tested individuals in a state (P(test + |tested)). For untested individuals who had mild or no symptoms, we assumed that the probability they tested positive was 0.2% to 32% P(test + |tested). Accordingly, we defined *α* such that *α*_0_ < P(test + |*S_1_*, untested)/P(test +|tested) < *α*_1_, with *α*_0_ = 80% and *α*_1_ = 99% and *β* such that *β*_0_< P(test + |*S*_0_ untested)/P(test +| tested) < *β*_1_, with *β*_0_ = 0.2% and *β*_1_ = 32%, where the distribution of *α* is truncated to lie within the interval (*α*_0_, *α*_1_) (likewise for *β*).

We calculated P(test+ | tested) as the cumulative number of cases divided by the cumulative number of tests in each state by April 18, 2020. We chose not to vary this quantity by date because the low testing rates per state resulted in unstable estimates of P(test+ | tested), particularly when less than 1% of the population was tested, which was the case in most states until early-mid April (Appendix Figure 3). Furthermore, when restricting to dates when at least 0.6% of the population was tested in each state, P(test+ | tested) was relatively stable within each state over time, suggesting that at least over the period considered, this quantity did not change significantly over time (Appendix Figure 4).

Because P(test + |tested) varied by state, P(test + |*S_1_*, untested) and P(test + |*S_0_*, untested) also varied by state. Otherwise, priors were the same across all states.

### Correction for imperfect test accuracy

To estimate the number of infections (*N*\*) correcting for imperfect test accuracy (i.e., sensitivity (*Se*) or specificity (*Sp*) <1), we used the following formula:^28^

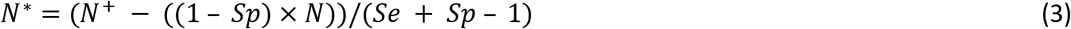

where *N*^+^ is the sum of the number of confirmed cases 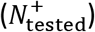 and the estimated number of infections among untested individuals correcting for incomplete testing 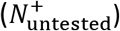.

### Probabilistic bias analysis

To account for uncertainty in our assumed priors, we used a probabilistic bias analysis.^21^ This method is semi-Bayesian because it defines prior distributions for bias parameters but not for the outcome (the estimated number of SARS-CoV-2 infections).^21^ We defined prior distributions for P(*S_1_*|tested), P(*S_1_*|untested), *α*, *β* P(*S_0_*|test +), SARS-CoV-2 test sensitivity, and SARS-CoV-2 test specificity based on available evidence (Table 1, Appendix Figure 1, Appendix Table 2).

#### Distribution of P(S_1_|tested)

We defined the distribution of P(S_1_|tested) as a truncated beta distribution with a range from 56-100% (median: 95%) under the assumption that the vast majority of individuals tested in the U.S. in March 2020 had moderate to severe symptoms. March 4, 2020, the CDC instructed physicians to use their judgment to determine which patients to test for SARS-CoV-2. Given the limited availability of tests in the U.S., they advised that symptomatic patients be prioritized for testing. On March 24, 2020, the CDC recommended that testing be prioritized for hospitalized patients, symptomatic healthcare workers, patients in vulnerable populations, and individuals with mild symptoms in communities experiencing a large number of COVID-19 hospitalizations.^22^ State-level testing priorities mostly followed CDC recommendations (Appendix Table 1).

#### Distribution of P(S_1_|untested)

We defined the distribution of P(*S*_1_ | untested) as a truncated beta distribution with a range from 0.2-15% (median: 5%). A study in Iceland that tested a random sample of individuals for SARS-CoV-2 reported that 0.9% individuals reported fever, 3.8% reported cough, and 0.6% reported a loss of smell or taste.^29^ While not directly comparable, studies of influenza symptoms, which overlap with COVID-19 symptoms, provide an estimate of the general frequency of fever and cough in the population. Since the majority of the population tested for SARS-CoV-2 in the U.S. had moderate to severe symptoms, these probabilities provide a reasonable estimate of P(*S_1_* |untested). Cohort studies of influenza-like illness (fever and cough and/or sore throat) in prior years in the U.S. and the Netherlands have found weekly incidence of 1-4%.^30-32^ Studies measuring over the entire influenza season have reported cumulative incidence of influenza-like illness ranging from approximately 7% to 17%.^33-35^

#### Distribution of α

To allow for state-level variation in P(test + |*S_1_*, untested) due to differing transmission dynamics in each state, our model allowed this probability to vary by state. We defined *α* as a truncated beta distribution ranging from 80% to 99% (median: 91%) such that *α_0_* < P(test + |*S*_1_, untested)/P(test + | tested) < *α*_1_. We used an empirical state-specific estimate of P(test+ | tested) equal to the cumulative number of cases divided by the cumulative number of tests by April 18, 2020. The median of state-specific distributions of P(test+|*S_1_*, untested) ranged from 0.02 to 0.46, and in many states the medians were close to 0.1 (Appendix Table 2). This is consistent with a study in Iceland, which estimated that P(test+|*S_1_*) was 1% among those tested in population screening (N=3,579) and 2% among those tested in a random sample (N=271).^29^

#### Distribution of β

To allow for state-level variation in P(test + | *S_0_*, untested) due to differing transmission dynamics in each state, our model allowed this probability to vary by state. We defined *β* as a truncated beta distribution ranging from 0.2% to 32% (median: 5%) such that *β*_0_ < P(test + | *S_0_*, untested)/P(test + | tested) < *β*_1_. As described above, we used an empirical state-specific estimate of P(test +| tested). The median of state-specific distributions of P(test+| *S_0_*, untested) ranged from 0.001 to 0.023 (Appendix Table 2). A study in Iceland estimated that P(test+| *S_0_*) was 0.8% among mild symptomatic/asymptomatic individuals tested in population screening and 0.6% (N=10,797) among mild symptomatic/ asymptomatic individuals tested in a random sample (N=2,283).^29^ One study in New York City screening 215 pregnant women admitted for delivery, regardless of symptoms, found that 13.7% of women without symptoms tested positive.^36^ However, these estimates may not be representative of the general population because of differences in age and immunity of pregnant women. The upper bound of P(test + |*S_0_*, untested) in states with high test positive probabilities includes this percentage (Appendix Table 2).

#### Distribution of P(S_0_|test +)

We defined the distribution of P(*S_0_*|test +) as a truncated beta distribution with a range from 25-70% (median: 49%) because studies that have tested both symptomatic and asymptomatic individuals have found estimated probabilities within this range.^2-4,6,29,37^ In these studies, it is possible that individuals who were asymptomatic at the time of testing developed symptoms later. Two other studies provide estimates from less generalizable populations. A study that tested the majority of passengers on the Diamond Princess cruise ship found that 17.9% of individuals who tested positive were asymptomatic, but the study population and transmission dynamics on the ship are not representative of the general U.S.^38^ One study in New York City screening 215 pregnant women admitted for delivery, regardless of symptoms, found that 87.9% of those who tested positive (N=33) were asymptomatic.^36^ However, we did not extend the maximum of our prior distribution to include this estimate because the study population is less representative than those of other studies estimating this quantity.

#### Distribution of SARS-CoV-2 test sensitivity

We defined the distribution of SARS-CoV-2 test sensitivity as a truncated beta distribution with a range from 65-100% (median: 87%) because the available evidence to date has reported clinical sensitivity within this range. The U.S. CDC RT-PCR test has analytical sensitivity ≥95% for RNA concentrations ≥1 copy/μL.^39^ A study of 213 RT-PCR-confirmed COVID-19 patients from Wuhan reported that the detection rate in the first 14 days since onset was 54-73% for nasal swabs and 30-61% for throat swabs.^10^ Using any RT-PCR as gold standard, another study of 1,014 patients from three Chinese provinces found that initial RT-PCR pharyngeal sensitivity ranged from 65-80%.^12^ Another small study (N=4) of COVID-19 patients from Wuhan also found that some that had been discharged tested positive 5-13 days later, suggesting that initial tests produced false negatives.^13^

#### Distribution of SARS-CoV-2 test specificity

We defined the distribution of SARS-CoV-2 test specificity as a truncated beta distribution with a range from 99.98%-100% (median: 99.90%). The SARS-CoV-2 RT-PCR diagnostic panel developed by the CDC was designed to minimize the chance of a false positive, so test specificity assumed to be very high in laboratories that comply with standard best practices, such as the use of negative controls.^39^ Validation of both the CDC and WHO real-time polymerase chain reaction (PCR) tests for SARS-CoV-2 found no false positives in cell culture samples containing other respiratory viruses.^27,40^ In one clinical evaluation of diagnostic accuracy, PCR testing had a very high specificity for SARS-CoV-2.^41^

### Constraining of prior distributions

We considered estimates of P(*S_0_*|test +) to be more reliable than those of P(test +|*S_1_*) and P(test + | *S_0_*) because P(*S_0_*|test +) has been estimated in published studies with more representative sampling.^2-4,37^ However, *φ* = P(*S_0_*|test +) is related to the parameters *θ* = {P(*S_1_* |untested), *α*, *β*} by a function (*M*: *θ* → *φ*) given by:

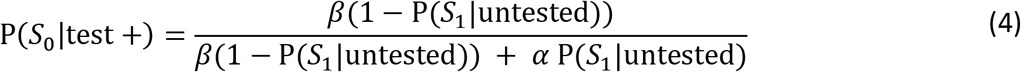

Mathematically, once a distribution is assigned to *θ* and *M* is defined, the distribution of *φ* is fully specified. However, because of the availability of reliable data on the distribution of plausible values for P(*S_0_*|test +), we used Bayesian melding^22^, a method which combines prior distributions on function inputs *θ* and output *φ* to generate a joint “melded” prior distribution on {*θ, φ*}. The advantage of this method is that it incorporates all available prior uncertainty as well as constraints on plausible output given by *M*. To sample values of {*θ, φ*} to be used as input to the probabilistic bias correction, we sampled 10^5^ variates from the melded prior distribution using a Sampling-Importance-Resampling algorithm.^22^

### Probabilistic bias correction

To estimate the number of SARS-CoV-2 infections accounting for uncertainty in our prior distributions, we sampled from the joint prior distributions and estimated the number of SARS-CoV-2 infections as described above using these randomly sampled prior values. We repeated this process 1,000 times to obtain a distribution of the expected case counts each day in each state. We used the median of the distribution of expected COVID-19 cases in each state on each day as the primary estimate. To assess uncertainty in our model, we report the simulation interval (2.5^th^ and 97.5^th^ percentiles of the distribution of total infections).

We calculated the percentage of the difference between observed case counts and the estimated number of infections attributable to imperfect test accuracy as the difference between *N*\* with *Se* and *Sp* set to our prior values and with *Se* = 1 and *Sp* = 1 divided by the difference between *N*\* and the observed case counts. We calculated the percentage attributable to incomplete testing as the 1 - the percentage attributable to imperfect test accuracy. To obtain national estimates of these percentages, we obtained the median and 2.5^th^ and 97.5^th^ percentiles of the state-specific distributions and weighted by state population.

## Data Availability

All data necessary to reproduce the findings of this study have been made publicly available at the GitHub repository https://github.com/jadebc/covid19-infections

## Code Availability

All code necessary to reproduce the entirety of findings of this study have been made publicly available at the GitHub repository https://github.com/jadebc/covid19-infections and a permanent archive of the version of the code that accompanied submission of this manuscript has been made at https://github.com/jadebc/covid19-infections/releases/tag/InitialSubmission

The file “README.md” in the root directory of the repository contains instructions on how to recreate all presented figures and tables in the manuscript.

## Author contributions

**Conceptualization:** Jade Benjamin-Chung

**Data Curation:** Anna Nguyen, Nolan N. Pokpongkiat, Stephanie Djajadi

**Formal Analysis:** Jade Benjamin-Chung, Anna Nguyen, Sean L. Wu

**Funding Acquisition:** Jade Benjamin-Chung

**Investigation:** Jade Benjamin-Chung, Yoshika S. Crider, Andrew Mertens

**Methodology:** Jade Benjamin-Chung, Anna Nguyen, Sean L. Wu, Alan Hubbard

**Project Administration:** Jade Benjamin-Chung

**Software:** Anna Nguyen, Nolan N. Pokpongkiat, Stephanie Djajadi, Anmol Seth, Andrew Mertens, Sean L. Wu

**Supervision:** Jade Benjamin-Chung

**Validation:** Anna Nguyen, Andrew Mertens

**Visualization:** Jade Benjamin-Chung, Anna Nguyen, Nolan N. Pokpongkiat, Stephanie Djajadi, Anmol Seth

**Writing – Original Draft Preparation:** Jade Benjamin-Chung, Sean L. Wu

**Writing – Review & Editing:** Jade Benjamin-Chung, Sean Wu, Yoshika S. Crider, Andrew Mertens, Michelle S. Hsiang, John M. Colford Jr., Art Reingold, Benjamin F. Arnold, Alan Hubbard

## APPENDIX

**Appendix Table 1.**
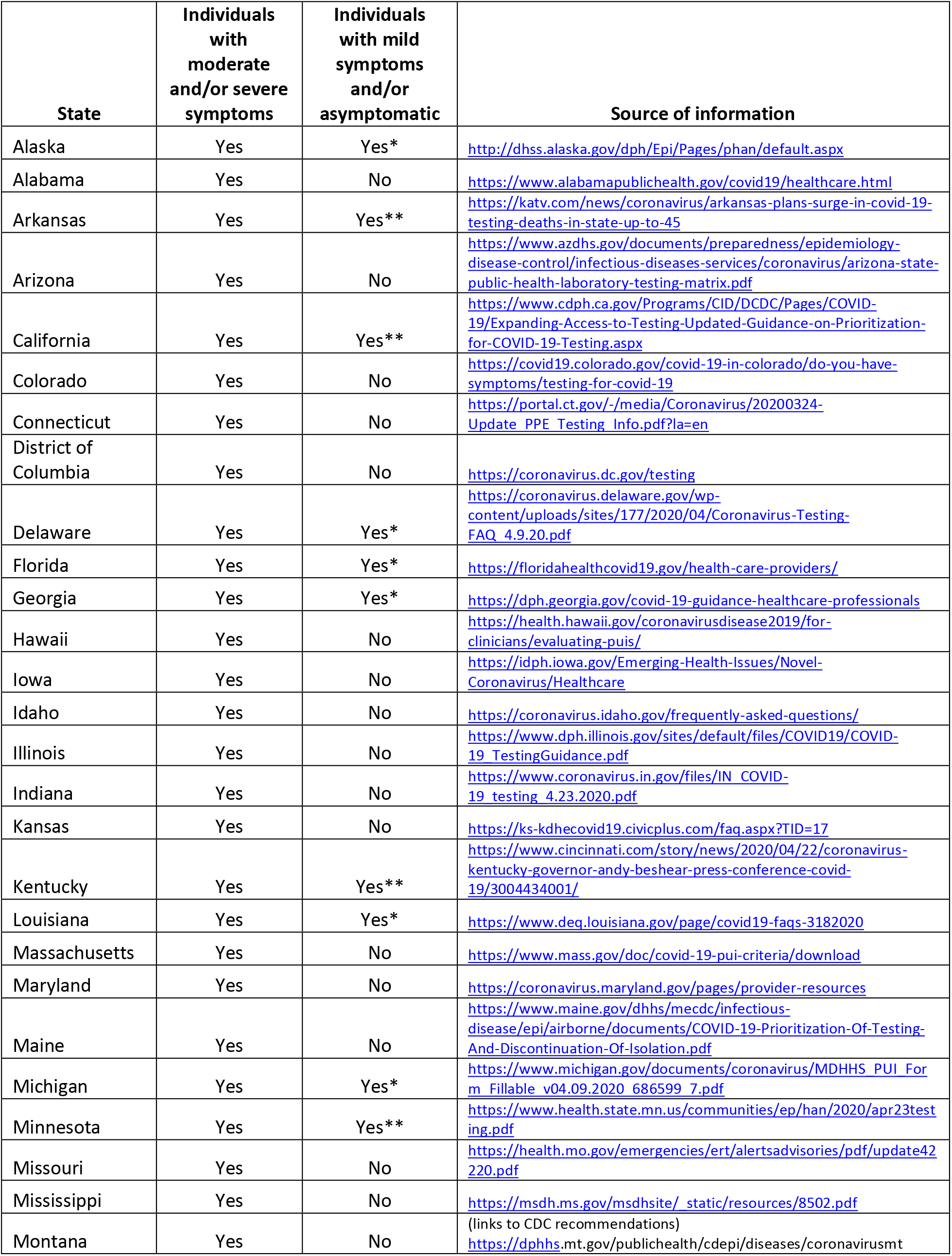
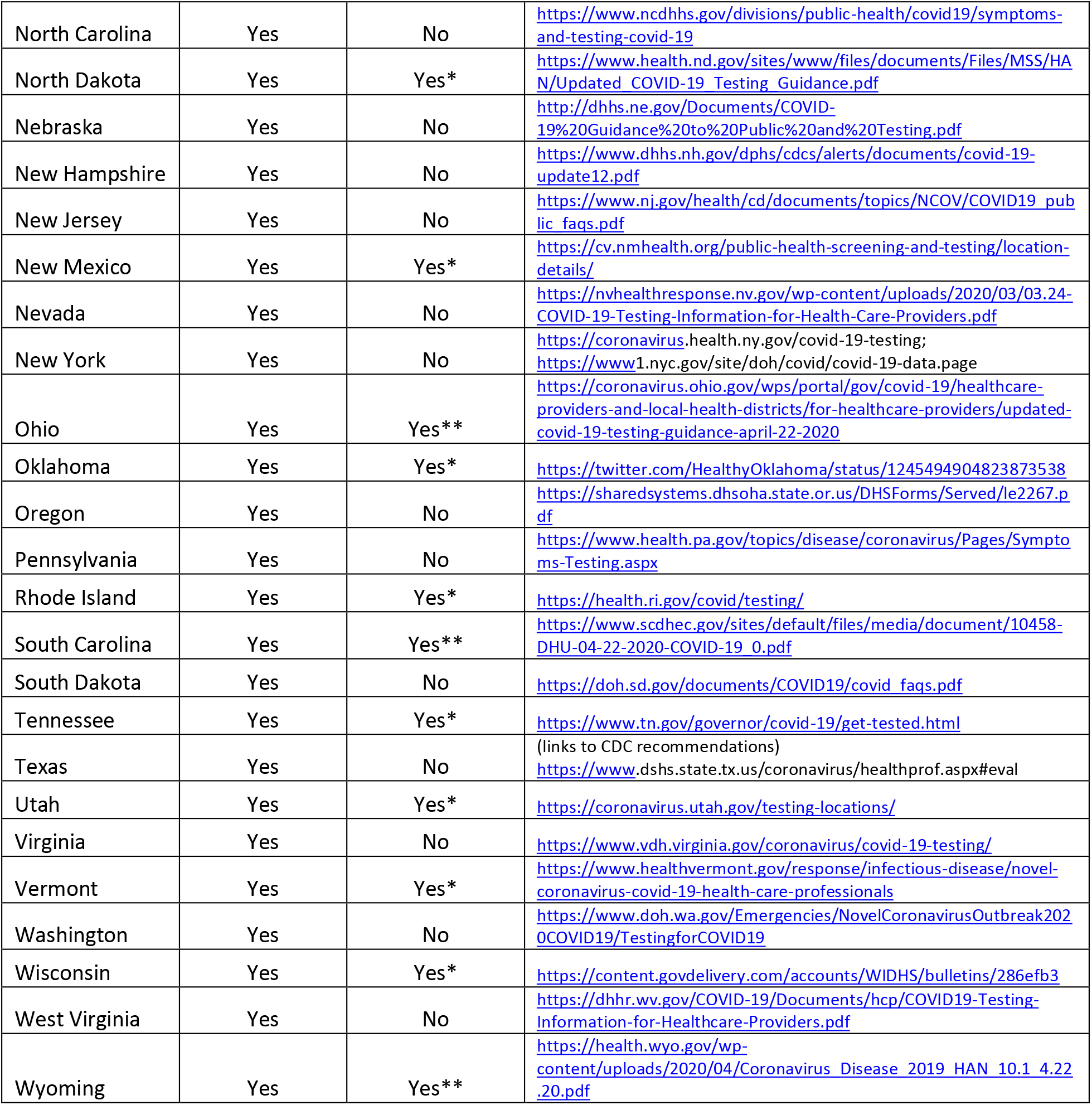
State-level recommendations for SARS-CoV-2 testing in the general population by symptom status

**Appendix Table 2.** State-specific prior distributions for probabilistic bias analysis

| State | P(test+ \| $S_1$) | | | P(test+ \| $S_0$) | | |
| --- | --- | --- | --- | --- | --- | --- |
|  | Minimum | Median | Maximum | Minimum | Median | Maximum |
| Alabama | 0.088 | 0.099 | 0.108 | 0.000 | 0.005 | 0.035 |
| Alaska | 0.026 | 0.03 | 0.032 | 0.000 | 0.001 | 0.01 |
| Arizona | 0.074 | 0.084 | 0.091 | 0.000 | 0.004 | 0.029 |
| Arkansas | 0.058 | 0.065 | 0.071 | 0.000 | 0.003 | 0.023 |
| California | 0.087 | 0.099 | 0.107 | 0.000 | 0.005 | 0.034 |
| Colorado | 0.167 | 0.19 | 0.206 | 0.001 | 0.01 | 0.066 |
| Connecticut | 0.242 | 0.275 | 0.299 | 0.001 | 0.014 | 0.096 |
| Delaware | 0.133 | 0.151 | 0.164 | 0.000 | 0.008 | 0.053 |
| District of Columbia | 0.161 | 0.183 | 0.199 | 0.001 | 0.009 | 0.064 |
| Florida | 0.082 | 0.093 | 0.101 | 0.000 | 0.005 | 0.033 |
| Georgia | 0.190 | 0.216 | 0.235 | 0.001 | 0.011 | 0.076 |
| Hawaii | 0.020 | 0.022 | 0.024 | 0.000 | 0.001 | 0.008 |
| Idaho | 0.080 | 0.091 | 0.098 | 0.000 | 0.005 | 0.032 |
| Illinois | 0.170 | 0.193 | 0.210 | 0.001 | 0.010 | 0.067 |
| Indiana | 0.150 | 0.17 | 0.185 | 0.000 | 0.009 | 0.059 |
| Iowa | 0.088 | 0.099 | 0.108 | 0.000 | 0.005 | 0.035 |
| Kansas | 0.081 | 0.092 | 0.100 | 0.000 | 0.005 | 0.032 |
| Kentucky | 0.066 | 0.075 | 0.081 | 0.000 | 0.004 | 0.026 |
| Louisiana | 0.137 | 0.155 | 0.169 | 0.000 | 0.008 | 0.054 |
| Maine | 0.045 | 0.052 | 0.056 | 0.000 | 0.003 | 0.018 |
| Maryland | 0.151 | 0.171 | 0.186 | 0.000 | 0.009 | 0.060 |
| Massachusetts | 0.186 | 0.211 | 0.229 | 0.001 | 0.011 | 0.074 |
| Michigan | 0.247 | 0.281 | 0.305 | 0.001 | 0.014 | 0.098 |
| Minnesota | 0.040 | 0.045 | 0.049 | 0.000 | 0.002 | 0.016 |
| Mississippi | 0.082 | 0.093 | 0.101 | 0.000 | 0.005 | 0.033 |
| Missouri | 0.082 | 0.093 | 0.101 | 0.000 | 0.005 | 0.033 |
| Montana | 0.032 | 0.037 | 0.04 | 0.000 | 0.002 | 0.013 |
| Nebraska | 0.067 | 0.076 | 0.082 | 0.000 | 0.004 | 0.026 |
| Nevada | 0.094 | 0.107 | 0.117 | 0.000 | 0.005 | 0.037 |
| New Hampshire | 0.079 | 0.090 | 0.098 | 0.000 | 0.005 | 0.031 |
| New Jersey | 0.401 | 0.455 | 0.495 | 0.001 | 0.023 | 0.159 |
| New Mexico | 0.038 | 0.044 | 0.047 | 0.000 | 0.002 | 0.015 |
| New York | 0.317 | 0.361 | 0.392 | 0.001 | 0.018 | 0.126 |
| North Carolina | 0.064 | 0.073 | 0.080 | 0.000 | 0.004 | 0.026 |
| North Dakota | 0.033 | 0.037 | 0.040 | 0.000 | 0.002 | 0.013 |
| Ohio | 0.101 | 0.115 | 0.125 | 0.000 | 0.006 | 0.040 |
| Oklahoma | 0.058 | 0.066 | 0.071 | 0.000 | 0.003 | 0.023 |
| Oregon | 0.039 | 0.045 | 0.048 | 0.000 | 0.002 | 0.016 |
| Pennsylvania | 0.161 | 0.183 | 0.199 | 0.001 | 0.009 | 0.064 |
| Rhode Island | 0.109 | 0.124 | 0.135 | 0.000 | 0.006 | 0.043 |
| South Carolina | 0.087 | 0.099 | 0.108 | 0.000 | 0.005 | 0.035 |
| South Dakota | 0.106 | 0.120 | 0.131 | 0.000 | 0.006 | 0.042 |
| Tennessee | 0.060 | 0.068 | 0.074 | 0.000 | 0.003 | 0.024 |
| Texas | 0.083 | 0.094 | 0.102 | 0.000 | 0.005 | 0.033 |
| Utah | 0.039 | 0.044 | 0.048 | 0.000 | 0.002 | 0.016 |
| Vermont | 0.051 | 0.058 | 0.063 | 0.000 | 0.003 | 0.020 |
| Virginia | 0.123 | 0.140 | 0.152 | 0.000 | 0.007 | 0.049 |
| Washington | 0.072 | 0.081 | 0.089 | 0.000 | 0.004 | 0.028 |
| West Virginia | 0.032 | 0.037 | 0.040 | 0.000 | 0.002 | 0.013 |
| Wisconsin | 0.069 | 0.079 | 0.086 | 0.000 | 0.004 | 0.028 |
| Wyoming | 0.037 | 0.042 | 0.046 | 0.000 | 0.002 | 0.015 |

**Appendix Table 3.**
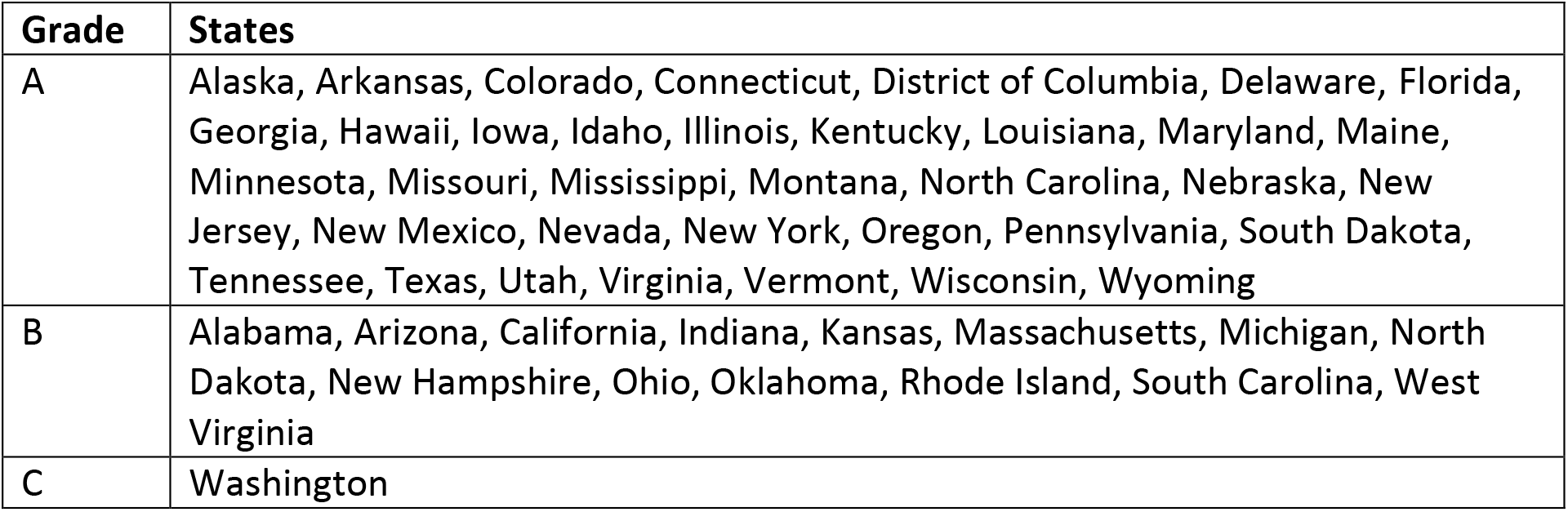
Quality of state-level test reports according to COVID Tracking Project

**Appendix Figure 1.**
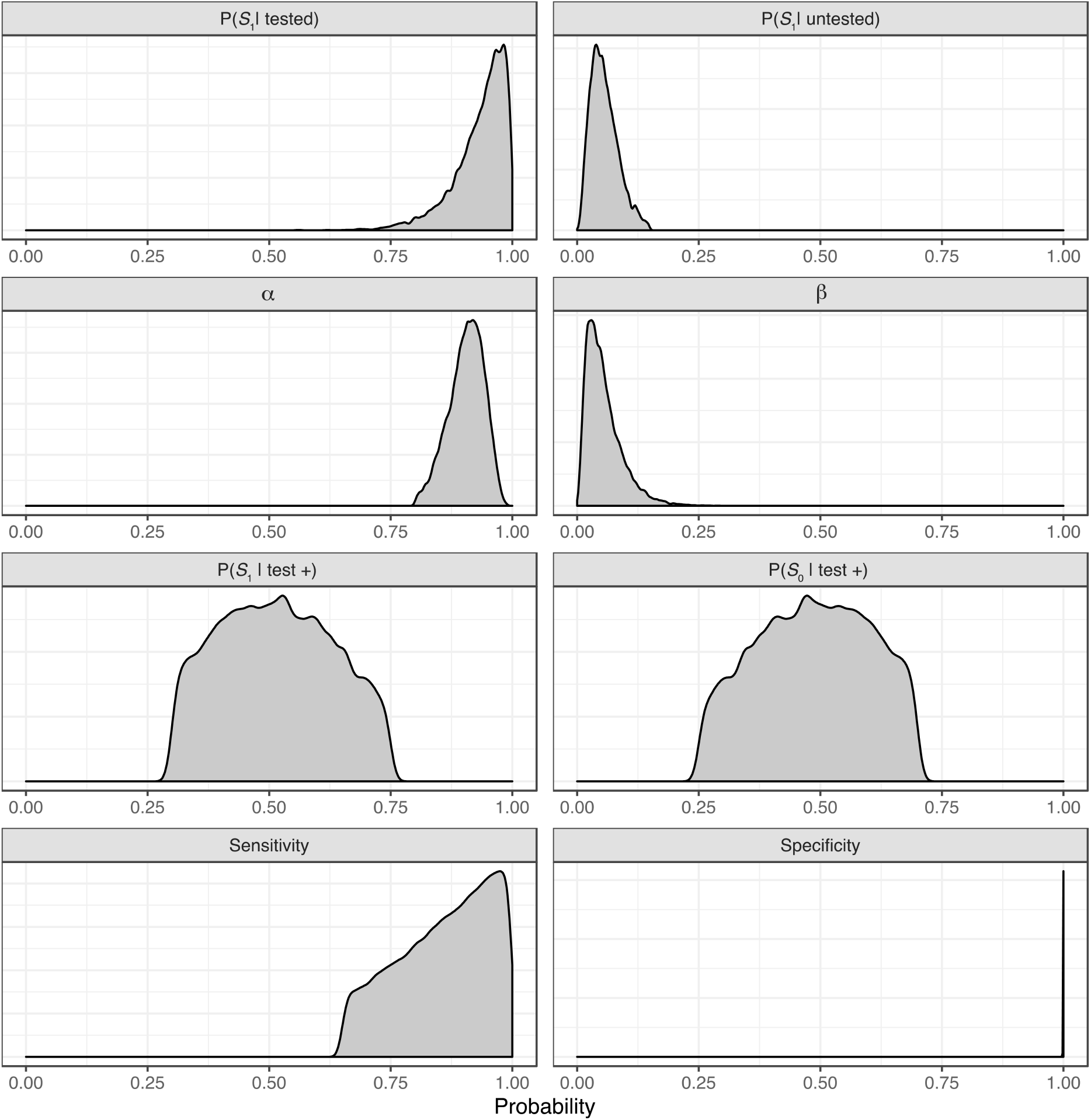
Static prior distributions for probabilistic bias analysis.

**Appendix Figure 2.**
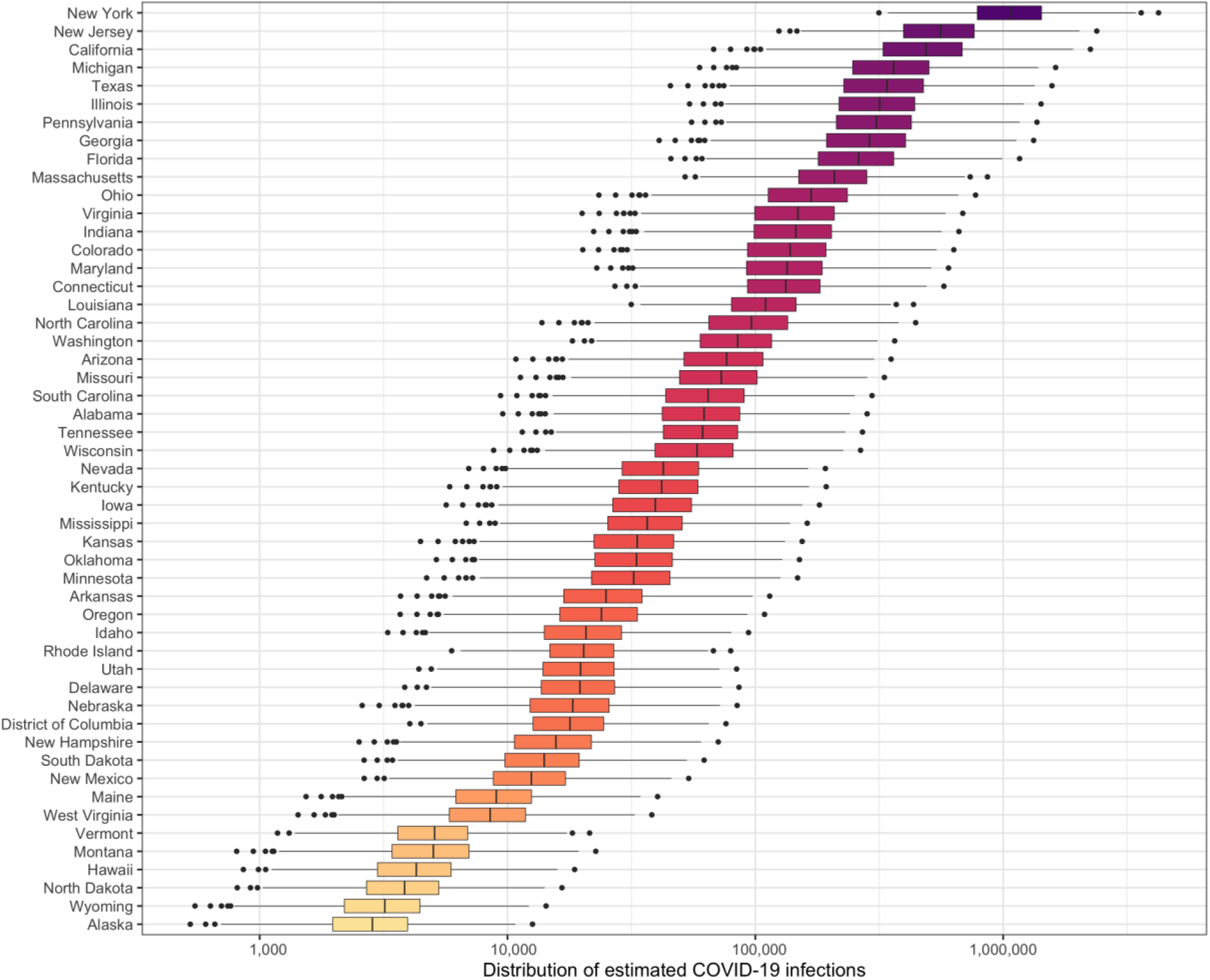
Distribution of expected SARS-CoV-2 infections by state correcting for bias due to incomplete testing and imperfect test accuracy.

**Appendix Figure 3.**
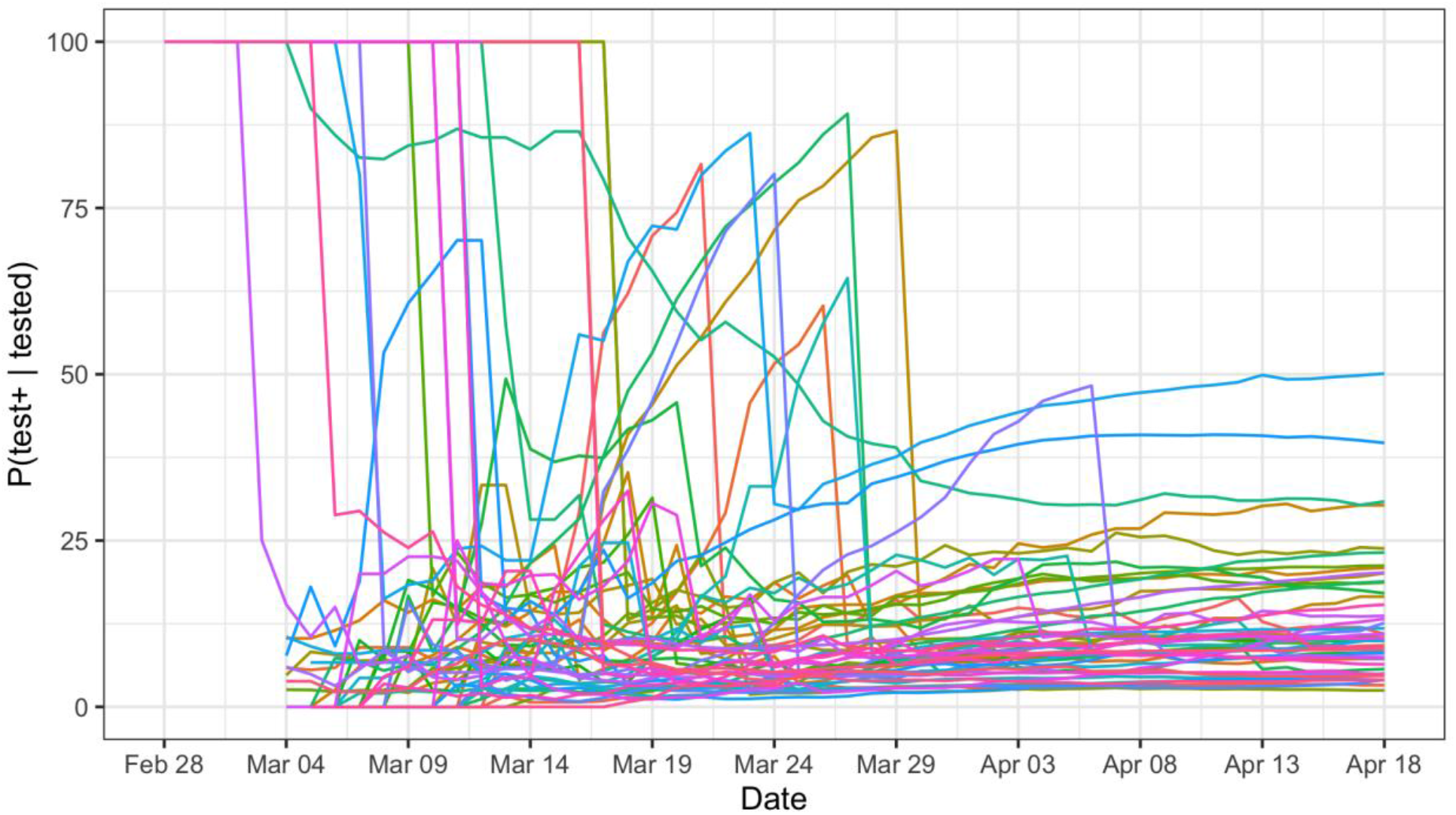
Probability of testing positive among those tested in each state by date.

**Appendix Figure 4.**
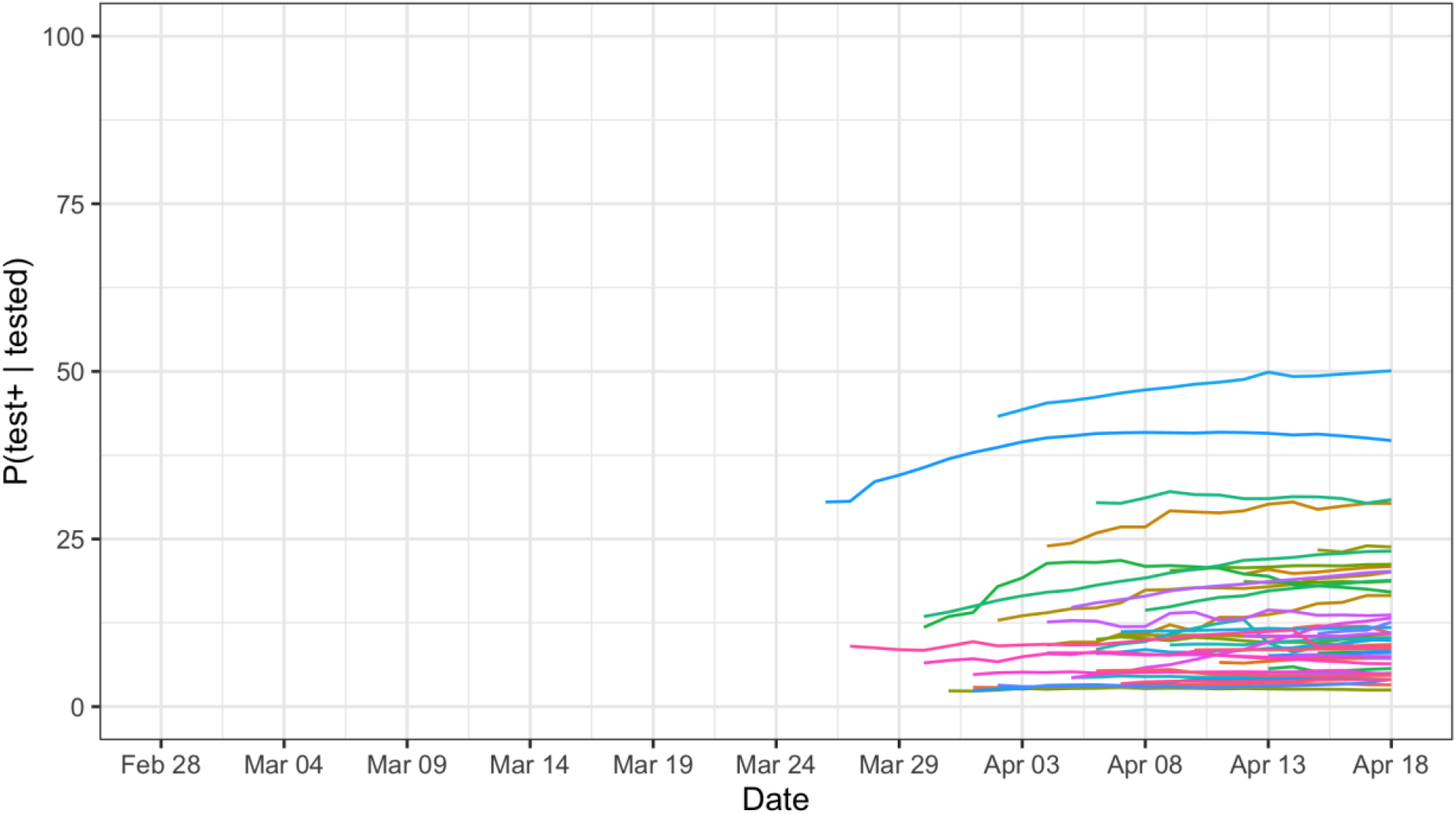
Probability of testing positive among those tested in each state by date restricting to dates when the percentage of the population tested was at least 0.6%.

## References

1 Lipsitch M, Swerdlow DL, Finelli L. Defining the Epidemiology of Covid-19 — Studies Needed. New England Journal of Medicine 2020; 382: 1194–6.

2 Day M. Covid-19: identifying and isolating asymptomatic people helped eliminate virus in Italian village. BMJ 2020; 368. DOI:10.1136/bmj.m1165.

3 Nishiura H, Kobayashi T, Miyama T, et al. Estimation of the asymptomatic ratio of novel coronavirus infections (COVID-19). *medRxiv* 2020;: 2020.02.03.20020248.

4 Wang C, Liu L, Hao X, et al. Evolving Epidemiology and Impact of Non-pharmaceutical Interventions on the Outbreak of Coronavirus Disease 2019 in Wuhan, China. *medRxiv* 2020;: 2020.03.03.20030593.

5 Qiu H, Wu J, Hong L, Luo Y, Song Q, Chen D. Clinical and epidemiological features of 36 children with coronavirus disease 2019 (COVID-19) in Zhejiang, China: an observational cohort study. The Lancet Infectious Diseases 2020; published online March 25. DOI:10.1016/S1473-3099(20)30198-5.

6 Lavezzo E, Franchin E, Ciavarella C, et al. Suppression of COVID-19 outbreak in the municipality of Vo, Italy. *medRxiv* 2020;: 2020.04.17.20053157.

7 Zou L, Ruan F, Huang M, et al. SARS-CoV-2 Viral Load in Upper Respiratory Specimens of Infected Patients. New England Journal of Medicine 2020; 382: 1177–9.

8 Du Z, Xu X, Wu Y, Wang L, Cowling BJ, Meyers LA. Early Release - Serial Interval of COVID-19 among Publicly Reported Confirmed Cases - Volume 26, Number 6—June 2020 - Emerging Infectious Diseases journal - CDC. 2020. DOI:10.3201/eid2606.200357.

9 Rothe C, Schunk M, Sothmann P, et al. Transmission of 2019-nCoV Infection from an Asymptomatic Contact in Germany. New England Journal of Medicine 2020; 382: 970–1.

10 Yang Y, Yang M, Shen C, et al. Evaluating the accuracy of different respiratory specimens in the laboratory diagnosis and monitoring the viral shedding of 2019-nCoV infections. *medRxiv* 2020;: 2020.02.11.20021493.

11 Wang W, Xu Y, Gao R, et al. Detection of SARS-CoV-2 in Different Types of Clinical Specimens. JAMA 2020; published online March 11. DOI:10.1001/jama.2020.3786.

12 Ai T, Yang Z, Hou H, et al. Correlation of Chest CT and RT-PCR Testing in Coronavirus Disease 2019 (COVID-19) in China: A Report of 1014 Cases. Radiology 2020;: 200642.

13 Lan L, Xu D, Ye G, et al. Positive RT-PCR Test Results in Patients Recovered From COVID-19. JAMA 2020; published online Feb 27. DOI:10.1001/jama.2020.2783.

14 Ferguson N, Laydon D, Nedjati Gilani G, et al. Report 9: Impact of non-pharmaceutical interventions (NPIs) to reduce COVID19 mortality and healthcare demand. 2020 DOI:https://doi.org/10.25561/77482.

15 Walker P, Whittaker C, Watson O, et al. The Global Impact of COVID-19 and Strategies for Mitigation and Suppression. 2020 https://www.imperial.ac.uk/media/imperial-college/medicine/sph/ide/gida-fellowships/Imperial-College-COVID19-Global-Impact-26-03-2020v2.pdf.

16 IHME COVID-19 health service utilization forecasting team, Murray CJ. Forecasting COVID-19 impact on hospital bed-days, ICU-days, ventilator-days and deaths by US state in the next 4 months. *medRxiv* 2020;: 2020.03.27.20043752.

17 Li R, Pei S, Chen B, et al. Substantial undocumented infection facilitates the rapid dissemination of novel coronavirus (SARS-CoV2). Science 2020; published online March 16. DOI:10.1126/science.abb3221.

18 Yue M, Clapham HE, Cook AR. Estimating the Size of a COVID-19 Epidemic from Surveillance Systems. Epidemiology 2020; Publish Ahead of Print. DOI:10.1097/EDE.0000000000001202.

19 Jewell NP, Lewnard JA, Jewell BL. Predictive Mathematical Models of the COVID-19 Pandemic: Underlying Principles and Value of Projections. JAMA 2020; published online April 16. DOI:10.1001/jama.2020.6585.

20 Jewell NP, Lewnard JA, Jewell BL. Caution Warranted: Using the Institute for Health Metrics and Evaluation Model for Predicting the Course of the COVID-19 Pandemic. Ann Intern Med 2020; published online April 14. DOI:10.7326/M20-1565.

21 Lash TL, Fox MP, Fink AK. Applying Quantitative Bias Analysis to Epidemiologic Data. Springer Science & Business Media, 2011.

22 Poole D, Raftery AE. Inference for Deterministic Simulation Models: The Bayesian Melding Approach. Journal of the American Statistical Association 2000; 95: 1244–55.

23 nytimes. nytimes/covid-19-data. The New York Times, 2020 https://github.com/nytimes/covid-19-data (accessed March 31, 2020).

24 Kissler SM, Tedijanto C, Goldstein E, Grad YH, Lipsitch M. Projecting the transmission dynamics of SARS-CoV-2 through the postpandemic period. Science 2020; published online April 14. DOI:10.1126/science.abb5793.

25 Salje H, Kiem CT, Lefrancq N, et al. Estimating the burden of SARS-CoV-2 in France. 2020; published online April 20. https://hal-pasteur.archives-ouvertes.fr/pasteur-02548181 (accessed April 21, 2020).

26 Modi C, Boehm V, Ferraro S, Stein G, Seljak U. Total COVID-19 Mortality in Italy: Excess Mortality and Age Dependence through Time-Series Analysis. *medRxiv* 2020;: 2020.04.15.20067074.

27 CDC. Interim Guidelines for Collecting, Handling, and Testing Clinical Specimens from Persons for Coronavirus Disease 2019 (COVID-19). Centers for Disease Control and Prevention. 2020; published online Feb 11. https://www.cdc.gov/coronavirus/2019-ncov/lab/guidelines-clinical-specimens.html (accessed April 23, 2020).

28 Rothman KJ, Greenland S, Lash TL. Modern Epidemiology. Philadelphia, PA: Lippincott Williams & Wilkins, 2008.

29 Gudbjartsson DF, Helgason A, Jonsson H, et al. Spread of SARS-CoV-2 in the Icelandic Population. New England Journal of Medicine 2020; In Press.

30 Chunara R, Goldstein E, Patterson-Lomba O, Brownstein JS. Estimating influenza attack rates in the United States using a participatory cohort. Scientific Reports 2015; 5: 1–5.

31 Patterson-Lomba O, Van Noort S, Cowling BJ, et al. Utilizing Syndromic Surveillance Data for Estimating Levels of Influenza Circulation. Am J Epidemiol 2014; 179: 1394–401.

32 Moberley S, Carlson S, Durrheim D, Dalton C. Flutracking: Weekly online community-based surveillance of influenza-like illness in Australia, 2017 Annual Report. Communicable Diseases Intelligence 2019; 43. DOI:https://doi.org/10.33321/cdi.2019.43.31.

33 Smolinski MS, Crawley AW, Baltrusaitis K, et al. Flu Near You: Crowdsourced Symptom Reporting Spanning 2 Influenza Seasons. Am J Public Health 2015; 105: 2124–30.

34 van Beek J, Veenhoven RH, Bruin JP, et al. Influenza-like Illness Incidence Is Not Reduced by Influenza Vaccination in a Cohort of Older Adults, Despite Effectively Reducing Laboratory-Confirmed Influenza Virus Infections. J Infect Dis 2017; 216: 415–24.

35 Friesema IHM, Koppeschaar CE, Donker GA, et al. Internet-based monitoring of influenza-like illness in the general population: Experience of five influenza seasons in the Netherlands. Vaccine 2009; 27: 6353–7.

36 Sutton D, Fuchs K, D’Alton M, Goffman D. Universal Screening for SARS-CoV-2 in Women Admitted for Delivery. New England Journal of Medicine 2020; DOI: 10.1056/NEJMc2009316

37 Dong Y, Mo X, Hu Y, et al. Epidemiological Characteristics of 2143 Pediatric Patients With 2019 Coronavirus Disease in China. Pediatrics 2020; published online March 1. DOI:10.1542/peds.2020-0702.

38 Mizumoto K, Kagaya K, Zarebski A, Chowell G. Estimating the asymptomatic proportion of coronavirus disease 2019 (COVID-19) cases on board the Diamond Princess cruise ship, Yokohama, Japan, 2020. Eurosurveillance 2020; 25: 2000180.

39 Centers for Disease Control and Prevention. CDC 2019-Novel Coronavirus (2019-nCoV) RealTime RT-PCR Diagnostic Panel. 2020 https://www.fda.gov/media/134922/download.

40 Corman VM, Landt O, Kaiser M, et al. Detection of 2019 novel coronavirus (2019-nCoV) by real-time RT-PCR. Eurosurveillance 2020; 25: 2000045.

41 Yam WC, Chan KH, Chow KH, et al. Clinical evaluation of real-time PCR assays for rapid diagnosis of SARS coronavirus during outbreak and post-epidemic periods. Journal of Clinical Virology 2005; 33: 19–24.

